# Absence of SARS-CoV-2 neutralizing activity in pre-pandemic sera from individuals with recent seasonal coronavirus infection

**DOI:** 10.1101/2020.10.08.20209650

**Authors:** Daniel Poston, Yiska Weisblum, Helen Wise, Kate Templeton, Sara Jenks, Theodora Hatziioannou, Paul Bieniasz

**Author notes:** Correspondence to: Paul D. Bieniasz, Laboratory of Retrovirology, The Rockefeller University, 31 1230 York Avenue, New York, NY, 10065., Theodora Hatziioannou, Laboratory of Retrovirology, The Rockefeller University, 31 1230 York Avenue, New York, NY, 10065.

## Abstract

Cross-reactive immune responses elicited by seasonal coronaviruses might impact SARS-CoV-2 susceptibility and disease outcomes. We measured neutralizing activity against SARS-CoV-2 in pre-pandemic sera from patients with prior PCR-confirmed seasonal coronavirus infection. While neutralizing activity against seasonal coronaviruses was detected in nearly all sera, cross-reactive neutralizing activity against SARS-CoV-2 was undetectable.

## Introduction

Since the initial description in December 2019 of a novel human coronavirus SARS-CoV-2, there has been a global effort to identify underlying the underlying causes for the great range of disease severity observed, from mild or even asymptomatic infection to severe respiratory distress and death. One hypothesis is that cross reactive immune responses, elicited by prior infection with seasonal coronaviruses impacts the course of SARS-CoV-2 infection, perhaps providing a degree of protection against severe COVID-19 disease.

The endemic seasonal human coronaviruses (HCoVs)—HCoV-HKU1, HCoV-OC43, HCoV-NL63, and HCoV-229E—cause, mild or subclinical respiratory infections, with severe disease being exceptionally rare [1]. Although there is low overall sequence homology between the SARS-CoV-2 Spike (S) protein and those of the endemic HCoVs, overlapping T-cell epitopes have been reported, particularly in the S2 subunit [2,3]. It is possible that neutralizing antibodies induced by seasonal HCoV infection could cross-react with similar epitopes in SARS-CoV-2 S. Such antibodies could potentially afford some level of protection against and perhaps contribute to the wide range of outcomes of SARS-CoV-2 infection. To investigate this possibility, we analyzed sera that had been collected prior to the COVID-19 pandemic from patients with a recent PCR-confirmed diagnosis of HCoV-OC43, HCoV-NL63, or HCoV-229E infection. Such samples should contain neutralizing antibodies against the respective seasonal HCoV, without the possibility of prior SARS-CoV-2 infection, allowing us to specifically test whether antibodies elicited by seasonal HCoV infection can neutralize SARS-CoV-2. Our results indicate a lack of SARS-CoV-2 cross-neutralization activity between the seasonal HCoVs and SARS-CoV-2.

## Methods

### Identification of Patient Samples

The thirty-seven prepandemic serum samples selected for inclusion in this study were all collected as part of routine clinical care prior to 2020 from patients in Edinburgh, Scotland, effectively excluding the possibility of prior SARS-CoV-2 infection. All samples were from symptomatic inpatients with PCR-confirmed diagnosis of HCoV-OC43, HCoV-NL63, or HCoV-229E infection, 11-291 days prior to collection of the serum sample. Ten positive control COVID-19 serum samples were collected in April-May 2020 from patients with mildly symptomatic, PCR-diagnosed SARS-CoV-2 infection, 24-61 days prior to serum collection. All samples were anonymized and ethical approval to utilize these patient samples was obtained through the NHS Lothian BioResource and the Rockefeller University IRB

### Viruses

The seasonal coronaviruses HCoV-OC43 (ATCC VR-759) and HCoV-229E (ATCC VR-740) were obtained from Zeptometrix, and HCoV-NL63 (Amsterdam I) was obtained from BEI resources. Viral stocks were generated by propagation on Huh7.5 cells. The replication-competent chimeric recombinant vesicular stomatitis virus encoding SARS-CoV-2 Spike and GFP (rVSV/SARS-2/GFP_2E1_) has been described previously and was propagated on 293T/ACE2cl.22 cells [4].

### Neutralization assays

Sera were initially diluted 1:12.5, and then serially diluted 5-fold over 7 dilutions in 96 well plates. Thereafter, approximately 4×10^3^ infectious units of either rVSV/SARS-2/GFP, HCoV-OC43, HCoV-NL63, or HCoV-229E were mixed with the serum dilutions and incubated at 37°C for 1 hour. Virus serum mixtures were subsequently transferred to 96-well plates containing 1×10^4^ 293T/ACE2cl.22 (for rVSV/SARS-2/GFP and HCoV-OC43) or HT1080/ACE2cl.14 (for HCoV-NL63 and HCoV-229E) target cells/well. Infection was allowed to proceed for 16 hours (rVSV/SARS-2) or 24 hours (HCoV-OC43, HCoV-NL63, HCoV-229E). The numbers of rVSV/SARS-2/GFP was assessed by flow cytometric detection of GFP expression as described previously [4]. For HCoV-OC43, HCoV-NL63, and HCoV-229E, cells were trypsinized and immunostained to detect nucleoprotein antigen expression in infected cells. For HCoV-OC43, Sigma MAB9013 was used, for HCoV-NL63, Eurofins M.30.HCo.B2D4 was used, for HCoV-229E: Eurofins M.30.HCo.B1E7 was used. A secondary antibody conjugate Alexa Fluor® 488 Goat anti-Mouse IgG (H+L) (Thermo) was then used to and infected cells enumerated by flow cytometry.

### Data analysis

All flow cytometry data was analyzed using FlowJo software version 10.6.1. All graphs and corresponding NT_50_ values were generated using GraphPad Prism version 8.

## Results

To assess whether prior infection by seasonal coronaviruses could elicit antibodies with neutralization activity against SARS-CoV-2, we identified 37 serum samples collected prior to the COVID19 pandemic from patients who were diagnosed using PCR with a seasonal coronavirus 11-291 days (median 80 = days) prior to serum sample collection. Of these 20 were diagnosed with HCoV-OC43 infection, 10 were diagnosed with HCoV-NL63 infection and 7 were diagnosed with HCoV-229E infection. We also developed flow cytometry-based coronavirus neutralization assays based on the detection of nucleocapsid expression in HCoV-OC43, HCoV-NL63, or HCoV-229E infected cells. Using theses neutralization assays, we confirmed that neutralizing antibodies targeting the seasonal coronaviruses were present in the pre-pandemic samples. Indeed, all sera from individuals diagnosed with recent infection by a seasonal coronavirus neutralized that same virus. Nevertheless, the neutralization titers varied between viruses. For example, while samples collected from HCoV-OC43 infected individuals typically exhibited potent neutralization of HCoV-OC43, sera collected from HCoV-229E infected individuals had comparatively weak neutralization activity against HCoV-229E. Most sera exhibited neutralizing activity against multiple seasonal coronaviruses. Indeed, some samples collected individuals with recent HCoV-229E infection neutralized HCoV-OC43 with higher titers than HCoV-229E. Collectively, 73% of samples had an NT_50_ > 500 for HCoV-OC43 and while 57% of samples NT_50_ > 500 for HCoV-229E, regardless of the virus detected at the time of sample collection. Neutralizing activity against HCoV-NL63 was typically of lower titer. Nevertheless all but one serum sample from individuals with recently diagnosed HCoV-NL63 infection had neutralizing activity against HCoV-NL63 with NT50 values of >1:50. Overall, this collection of serum samples had extensive neutralizing activity against several seasonal coronaviruses including, particularly, the betacoronavirus HCoV-OC43, that is the most closely related to SARS-CoV-2 of the viruses tested. Indeed, some of the sera had potent neutralizing activity against HCoV-OC43 with NT_50_ values in excess of 10,000.

In contrast, none of the very same 37 serum samples tested had any detectable neutralization activity against rVSV/SARS-CoV-2/GFP. Importantly, rVSV/SARS-CoV-2/GFP is at least as, or more, sensitive to neutralization by COVID-19 plasma analysis as SARS-CoV-2 [4]. Indeed, sera collected from ten individuals with recently diagnosed SARS-CoV-2 infection could neutralize rVSV/SARS-CoV-2/GFP with NT_50_ values ranging from 96 to 5400. Overall these data strongly suggest that only pandemic sera, and not pre-pandemic sera have neutralizing activity against SARS-CoV-2, and further suggest that pre-existing serological immunity to seasonal coronaviruses is not a major driver of the diverse outcome of SARS-CoV-2 infection.

## Discussion

These data demonstrate that neutralization activity against seasonal coronaviruses is nearly ubiquitous in sera collected from individuals with PCR-confirmed pre-pandemic seasonal coronavirus infection. Indeed, most sera had neutralizing activity against multiple seasonal coronaviruses and some sera had greater neutralization potency against different coronaviruses than the one detected at the time of sample collection. This may be due to inherent differences in neutralization sensitivity among the seasonal coronaviruses, and is most likely the result of prior, undocumented infection with different seasonal coronaviruses. That we observed more potent antibody responses to HCoV-OC43 and regardless of PCR result may suggest recent infection with this virus is more common, which is in line with previous observations suggesting that reinfection with HCoV-OC43 and HCoV-229E occurs at a greater frequency than HCoV-NL63[5–8] and that infection with HCoV-OC43 is common this geographic locale.

While we cannot exclude the possibility that that seasonal coronavirus elicit cross-neutralizing antibodies, the divergence between seasonal coronaviruses S proteins would suggest limited cross reactivity (HCoV-OC43 S shares 22.6 and 24.5% identical amino acids with and HCoV-229E and HCoV-NL63 S respectively, while HCoV-NL63 and HCoV-229E S share 55% identical amino acids) Accordingly, none of the samples tested had any neutralization activity against SARS-CoV-2 whose spike protein shares 24%-29% amino acid identity with the seasonal coronaviruses. In agreement with the notion that there is little cross reactivity between seasonal HCoV neutralizing antibodies and SARS-CoV-2, many of the monoclonal antibodies cloned from SARS-CoV-2 infected individuals contain very low levels of somatic hypermutation [9], suggesting that they arise from *de novo* rather than recall B-cell responses. However, instances of cross-reactive antibodies with high levels of somatic hypermutation have been reported, indicating that in some cases memory B cells evoked by prior seasonal HCoV infection may be recalled during infection with a SARS-like coronavirus [10].

While other groups have reported the existence of SARS-CoV-2 cross-reactive neutralizing antibodies in sera from individuals that were not infected SARS-CoV-2, the neutralization activity observed appears low [11,12]. Unlike other reports the pre-pandemic sera used in our study that have undetectable neutralization activity against SARS-CoV-2 can neutralize seasonal HCoVs, in some cases quite potently. While it is possible that there are rare instances of individuals possessing antibodies from prior seasonal HCoV infection may be able to also target SARS-CoV-2 S, our data argues against a broad role for pre-existing protective humoral immunity against SARS-CoV-2.

## Data Availability

All data available on reasonable request

## Author Contributions

HW, SJ, YW, TH, and PDB conceived and designed the study. DP performed the neutralization assays. DP, TH, and PDB wrote the manuscript with input from all authors.

---

**Figure 1.**
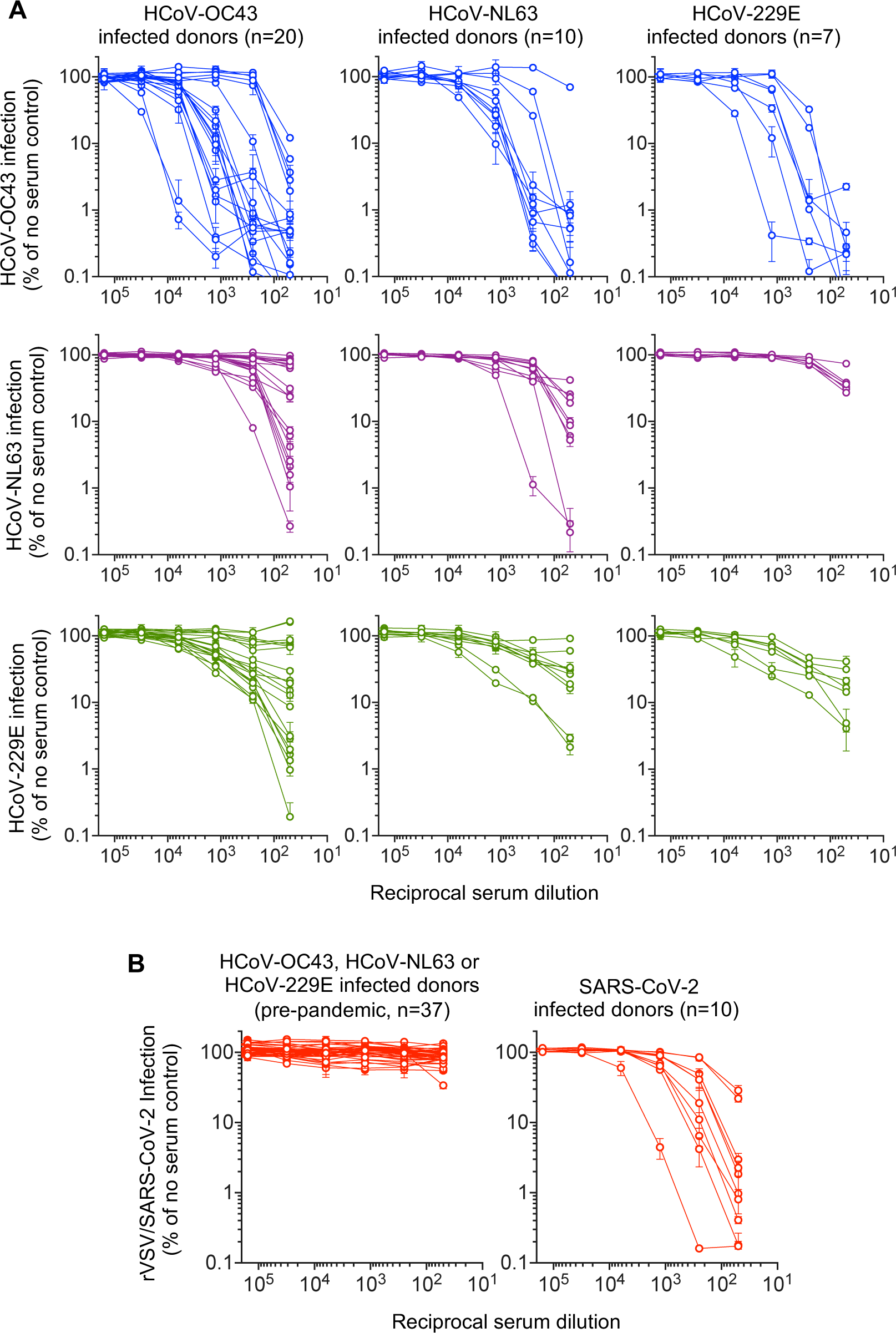
Coronavirus neutralizing activity in sera from individuals with recent coronavirus infection. **(A)** Infection by HCoV-OC43 (blue), HCoV-NL63 (purple) and HCoV-229E (green) in the presence of the indicated dilutions of pre-COVID-19-pandemic sera, from individuals recently diagnosed by PCR with HCoV-OC43, HCoV-NL63, or HCoV-229E infection, as indicated. Infected cells were enumerated by flow cytometry and the number of infected cells is plotted a percentage of the number of infected cells (∼30%) obtained in the absence of serum. **(B)** Infection by rVSV/SARS-CoV-2 in the presence of the indicated dilutions of pre-COVID-19-pandemic sera from individuals recently diagnosed by PCR with HCoV-OC43, HCoV-NL63, or HCoV-229E infection (left panel), or COVID-19 convalescent sera (right panel). Infected cells were enumerated by flow cytometry and the number of infected cells is plotted a percentage of the number of infected cells (∼30%) obtained in the absence of serum.

